# Different genetic liabilities to neuropsychiatric conditions in suicides with no prior suicidality

**DOI:** 10.1101/2025.05.02.25326877

**Authors:** Hilary Coon, Andrey A. Shabalin, Eric T. Monson, Emily DiBlasi, Seonggyun Han, Lisa M. Baird, Erin A. Kaufman, Doug Tharp, Michael J. Staley, Zhe Yu, Qingqin S. Li, Sarah M. Colbert, Amanda V. Bakian, Anna R. Docherty, Andrew M. McIntosh, Heather C. Whalley, Dierdre Amaro, David K. Crockett, Niamh Mullins, Brooks R. Keeshin

**Author notes:** corresponding author: Hilary Coon, University of Utah Department of Psychiatry, BPRB Room 408, 20 S 2030 East, Salt Lake City, UT 84112.

## Abstract

**Importance.:** Though suicide attempt is the most robust predictor of suicide death, few who attempt go on to die by suicide (<10%), and ∼50% of all suicide deaths occur in the absence of evidence of prior attempts. Risks in this latter group are particularly poorly understood.

**Objective.:** Data from the Utah Suicide Mortality Risk Study (USMRS) were used to study underlying polygenic liabilities among suicide deaths without evidence of prior nonfatal suicidal thoughts or behaviors (SD-N) compared to suicide deaths with prior nonfatal suicidality (SD-S).

**Design.:** We used an analysis of covariance design, comparing SD-N to SD-S and to population controls with similar genetic ancestry from the United Kingdom.

**Setting.:** We selected 12 source studies to generate descriptive quantitative polygenic scores (PGS) reflecting neuropsychiatric conditions. Analysis of covariance was used to evaluate suicide mortality subsets and controls adjusted for sex, age, and genetic ancestry effects.

**Participants.:** Suicide deaths were population-ascertained through a 25-year collaboration with the Utah State Office of the Medical Examiner. Evidence of suicidality was determined from diagnoses and clinical notes, yielding 1,364 SD-N and 1,467 SD-S deaths, compared to 20,368 controls.

**Main Outcomes.:** The tested PGS spanned 12 psychiatric, neurodevelopmental, and neurodegenerative conditions.

**Results.:** SD-N were significantly more male (82.33% vs. 67.76%) and older at death (47.26 years vs. 41.36 years) than SD-S. Controls were significantly less male than both suicide subsets (43.71%). Genetic ancestry was similar across suicide subsets and controls (% European: 96.77%, 96.81%, and 97.38%). Comparing SD-N to SD-S revealed significantly lower PGS in SD-N for: MDD (p=0.0015), neuroticism (p=0.0016), anxiety (p=0.0048), Alzheimer’s (p=0.011), depressed affect (p=0.015), schizophrenia (p=0.020), PTSD (p=0.023), and bipolar disorder (p=0.028). This attenuation in SD-N was particularly pronounced for depressed affect, neuroticism, and Alzheimer’s, where PGS were not different from controls. Sex-specific analyses suggested attenuation of PGS in SD-N was driven by males for MDD, anxiety, and PTSD, and by females for bipolar disorder, neuroticism, and Alzheimer’s.

**Conclusions and Relevance.:** SD-N have significantly different genetic liabilities from SD-S, particularly regarding neuropsychiatric conditions. Results have far-reaching implications both for future research and for preventions for those at highest risk of mortality.

**KEY POINTS:** 

**Question:** What are underlying genetic liabilities related to neuropsychiatric conditions in the roughly half of suicide deaths with no evidence of prior nonfatal suicidal thoughts or behaviors (SD-N), a group that has not previously been accessible for study?

**Findings:** These suicide deaths with no prior nonfatal suicidality showed significantly attenuated underlying polygenic liabilities associated with mental health traditionally thought to be core features of suicide mortality risk, and justifies additional studies of underlying risks associated with non-psychiatric conditions and behaviors.

**Meaning:** These differences in underlying liabilities between suicide deaths with and without prior suicidality suggest departure from the traditional mental health risks that have been the focus of suicide risk discovery, and impel new directions for future research and prevention efforts.

## INTRODUCTION

Suicide death is a significant public health crisis, with 49,449 deaths reported in 2022 in the U.S.^1^ and >700,000 per year worldwide.^2^ Risks specific to suicide mortality remain largely elusive.^3–4^ The most robust predictor of suicide mortality is a prior attempt, but few individuals who make suicide attempts go on to die by suicide, with estimates ranging from 2.5% - 7%.^5–7^ In addition, more than half of suicide deaths occur in the absence of evidence of prior suicidal thoughts or behaviors,^8–10^ and a more than half also show no evidence of psychiatric diagnoses.^11–13^ These results suggest that while prior attempts and psychopathology are important, they are insufficient predictors of suicide death.

Our prior study using Utah Suicide Mortality Risk Study (USRMS)^15^ resources showed that prevalence of 32 neuropsychiatric diagnoses was significantly attenuated among suicide deaths with no evidence of prior suicidality (SD-N) when compared to suicides with prior non-lethal suicidality (SD-S). Such results could be attributed to poor mental health screening, poor access to mental health care or to low care-seeking behaviors. However, results could also be due to differences in underlying liabilities to these disorders, a plausible hypothesis given the increasing body of evidence outlined above. USMRS data resources can address this knowledge gap. We have studied Utah suicides classified as either SD-N or SD-S, using polygenic scores (PGS) as proxies for underlying risks of 12 conditions spanning the neuropsychiatric spectrum. We calculated the PGS using summary statistics from 12 recent studies selected for size and for matching of ascertainment and genetic ancestry to the USMRS resource using a pre-registered study design.^16^

## METHODS

### Suicide death sample

This study used data from a subset of 4,251 suicide deaths from the USMRS sample with electronic health records, clinical notes for natural language processing (NLP), and genome-wide array genotyping data. The USMRS resource has been described in detail elsewhere.^17–18^ Briefly, biosamples were collected from suicide deaths through a long-term collaboration with the Utah State Office of the Medical Examiner (OME). Suicide determination was made by the OME following detailed investigation of the scene and circumstances of the death. Because the state of Utah has a single central OME, all deaths in this study were evaluated by the same office/processes, providing a consistent approach to suicide determination, an advantage over states with decentralized systems. When sufficient sample amounts were available, high quality DNA was extracted. Identifiers from suicide deaths were securely transferred to staff at the Utah Population Database (UPDB), a state-wide database that contains over 27 million data records on over 11 million individuals, including demographics and comprehensive health records data.^19^ UPDB staff linked health records and supplied clinical notes to an NLP algorithm that detects suicidality mentions (see below for details). For this study, identifiers were stripped, rare diagnoses and demographic groups were aggregated, and NLP results were collapsed into ratings of positive/negative suicidality before data were given to the research team. This study was approved by Institutional Review Boards from the University of Utah, Intermountain Health, and the Utah Department of Health and Human Services.

### Population controls

Control genotype data was used from population cohorts similar to the Utah suicide cohort regarding genetic ancestry, including data (N=20,368) from the UK10K resource^20^ and unrelated individuals from the Generation Scotland resource.^21^ Molecular data from these control resources represents broad population ascertainment, although the UK10K resource includes some enrichment for individuals with common and rare health conditions, and may therefore be somewhat conservative as a comparison group. Available control data consisted only of genetic information; assessment or exclusion of suicidality was not possible.

### Demographic and clinical data on suicide deaths

Demographic and clinical data defining the USMRS suicides included age at death, sex, and clinical notes and diagnoses from electronic health records. Suicides were categorized as having evidence of prior nonfatal suicidality based on diagnostic data through ICD-9 or ICD-10 diagnostic codes according to the National Center for Health Statistics definition.^22^ Diagnoses that occurred within one week prior to death were excluded so that diagnostic codes associated with the final fatal suicide attempt were not mis-attributed to prior nonfatal suicidality. Because suicidality is not well captured in diagnostic data, we supplemented the SD-N and SD-S subtype definition using data from a validated NLP algorithm to detect suicidality in clinical notes.^15^ We applied a broad definition of suicidality from this NLP such that the SD-S subtype comprised suicide deaths with any mention of prior suicidal ideation or behavior in order to make the SD-N as accurate as possible for absence of prior suicidality. As found previously,^15^ using this approach of incorporating the NLP information increased the number of SD-S by 32% above using a definition from diagnostic codes alone.

### Genotyping of suicide deaths

Genotyping on suicide deaths was done using the Illumina Infinium PsychArray,^23^ which includes 560,000 single nucleotide polymorphisms (SNPs). Genotyping quality control was performed using SNP clustering in the Illumina GenomeStudio software.^24^ SNPs were retained if the GenTrain score was >0.5 and the cluster separation score was >0.4. SNPs with >5% missing genotypes were removed; samples with a call rate <95% were removed. The SNPs assayed in both suicides and controls were combined and jointly imputed. Genetic ancestry was estimated with a modification of the kgp2anc algorithm^25^ using 1000Genomes data^26^ as a reference.

### Computation of polygenic scores (PGS)

Polygenic scores (PGS) were computed for SD-N (N=1,364), SD-S (N=1,467), and controls (N=20,368) using available summary statistics from 12 studies spanning the neuropsychiatric spectrum (Supplementary Table S1, source sample N’s ranged from 31,880-1,126,563). These recent, large studies were chosen to match to the USMRS regarding ancestry (primarily European) and ascertainment (population rather than military cohorts), and pre-registered as part of our study design.^16^ PRSice 2.0^27^ was used to calculate individual PGS. PGS are weighted quantitative summary scores reflecting genetic liability to a trait or diagnosis rather than the binary presence or absence of that trait. A score for an individual in the target sample is the summation of dosage of minor alleles for each SNP multiplied by the effect size of that allele at that SNP in the discovery GWAS. We did not apply any p-value cut-off to select SNPs from the source studies to be inclusive of all genetic data. PGS in this study are descriptive, broadly associated with underlying polygenic liabilities to the diagnoses and also any additional pleiotropic genetic liabilities.^28^

### Analyses

Analysis of covariance was used to compare SD-N (N=1,364) to SD-S (N=1,467) and each subset to controls (N=20,368), adjusting for age, sex, and 20 residual genetic ancestry principal components (PCs), and controlling for multiple testing through the Benjamini-Hochberg procedure (false discovery rate method).^14^ For ease of interpretation, PGS from the suicide mortality subsets for each PGS outcome were normalized to controls. Because many neuropsychiatric conditions have strong sex differences, tests were stratified by sex. Within males, there were 1,123 SD-N, 994 SD-S, and 8,902 controls. Suicide death is heavily dominated by males; therefore, the female suicide sample sizes were smaller, with 241 SD-N, 473 SD-S, and 11,466 controls. Within-sex analyses were performed as described above concerning covariates, multiple testing adjustment, and normalizing to controls.

## RESULTS

Table 1 shows descriptive characteristics of Utah suicide death subtypes and controls. Suicide deaths without evidence of prior nonfatal attempts (SD-N, N=1,364) included significantly more males with an older mean age at death compared to suicides with evidence of prior nonfatal attempts (SD-S, N=1,467). The two suicide death subtypes were not significantly different in terms of genetic ancestry; both were predominantly European. Matching population expectations,^1,2^ the UK ancestry-matched control data (N=20,333, 97.38% European ancestry) included significantly more females (56.29%) than either of the suicide death subtypes, which were both predominantly male.

**Table 1.** Demographic characteristics of suicide death subtypes.

| Characteristic | Both male and female suicide deaths |  |  | Male suicide deaths |  |  | Female suicide deaths |  |  |
| --- | --- | --- | --- | --- | --- | --- | --- | --- | --- |
|  | SD-N<br>(N=1,747) | SD-S<br>(N=2,253) | p-value | SD-N<br>(N=1,123) | SD-S<br>(N=994) | p-value | SD-N<br>(N=241) | SD-S<br>(N=473) | p-value |
| Mean age at death<br>(std. dev.) | 47.26<br>(18.92) | 41.36<br>(15.59) | 6.38E-17 | 47.78<br>(19.58) | 41.36<br>(16.10) | 2.92 E-15 | 44.79<br>(15.22) | 41.36<br>(14.48) | 0.00480 |
| Percent European<br>ancestry (std. dev.) | 96.77%<br>(2.93) | 96.81%<br>(2.96) | 0.718 (NS) | 96.81%<br>(2.86) | 96.77%<br>(3.11) | 0.716 (NS) | 96.56%<br>(3.27) | 96.89%<br>(2.63) | 0.145 (NS) |
| Percent male | 82.33% | 67.76% | 2.53E-13 |  |  |  |  |  |  |

NOTES: SD-N = suicide deaths with no evidence of prior nonfatal suicidality. SD-S = suicide deaths with evidence of prior nonfatal suicidality.
Percentage of European genetic ancestry was determined using data from the 1000 Genomes Project.
For comparison, population control data from the UK (UK10K and Gen Scotland data) was 97.38% European ancestry and 56.29% female.

### Polygenic score comparisons

Table 2 and Figure 1 show results of differences across the SD-N and SD-S subsets. For ease of interpretation, all results are normalized to the UK controls, giving the UK controls mean PGS scores of zero for each trait. Of note, PGS showed similar 95% confidence intervals across tested phenotypes (Figure 1). Results indicated significantly lower PGS in the SD-N vs. SD-S subtype comparison for bipolar disorder, major depressive disorder (MDD), depressed affect, anxiety, post-traumatic stress disorder (PTSD), neuroticism, schizophrenia, and Alzheimer’s disease (AD). Comparisons of the SD-N subtype to the UK controls showed that for depressed affect, neuroticism, and AD, the SD-N subtype was indistinguishable from the control standard mean of zero. In addition, though scores for autism, ADHD, and alcohol were not significantly different in the SD-N vs. SD-S test, they showed modestly significant and similar elevations over the control PGS in both SD-N and SD-S subtypes.

**Figure 1.**
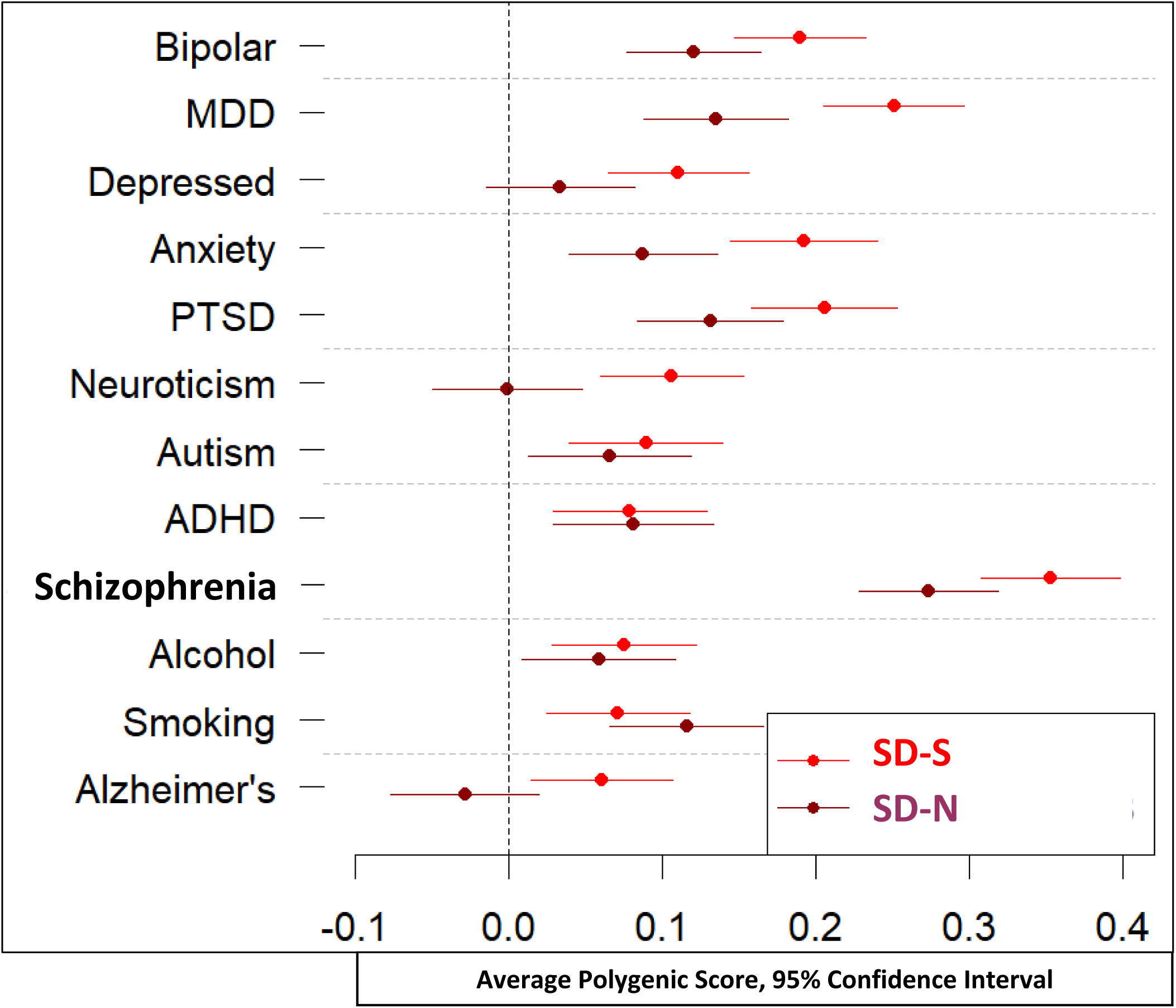
Polygenic score (PGS) analysis of covariance results for suicide mortality subtypes. Results for suicide subtypes are normalized to the UK controls, which are represented by the zero line in the graph. All tests were adjusted for sex, age, and 20 genetic ancestry principal components. SD-N = suicide deaths with no evidence of prior nonfatal suicidality. SD-S = suicide deaths with evidence of prior nonfatal suicidality. MDD = major depressive disorder. PTSD = post-traumatic stress disorder. ADHD = attention deficit hyperactivity disorder.

**Table 2.** Polygenic score (PGS) analysis of covariance results comparing suicide death subsets.

| Scored trait or diagnosis | SD-N mean PGS | SD-S mean PGS | SD-N vs. SD-S (p-value; FDR-adj p-value) | SD-N vs. UK Controls (p-value; FDR-adj p-value) | SD-S vs. UK Controls (p-value; FDR-adj p-value) |
| --- | --- | --- | --- | --- | --- |
| Bipolar disorder | 0.121 | 0.190 | **0.101 (0.0281; 0.0421 )** | **0.132 (4.95E-06; 1.20E-05)** | **0.200 (3.31E-13; 1.32E-12)** |
| MDD | 0.135 | 0.251 | **0.136 (0.00152; 0.0182)** | **0.150 (1.96E-07; 1.18E-06)** | **0.264 (9.16E-22; 5.50E-21)** |
| Depressed Affect | 0.034 | 0.111 | **0.103 (0.0147; 0.0353)** | 0.036 (NS) | **0.113 (3.81E-05; 7.63E-05)** |
| Anxiety | 0.088 | 0.192 | **0.117 (0.00477; 0.0191)** | **0.089 (0.00199; 0.00341)** | **0.194 (1.54E-12; 3.69E-12)** |
| PTSD | 0.131 | 0.206 | **0.096 (0.0228; 0.0391)** | **0.116 (5.01E-05; 1.20E-04)** | **0.195 (9.15E-13; 2.75E-12)** |
| Neuroticism | -0.001 | 0.106 | **0.132 (0.00163; 0.00976)** | -0.003 (NS) | **0.107 (8.71E-05; 1.49E-04)** |
| Autism | 0.066 | 0.089 | 0.021 (NS) | **0.068 (0.0168; 0.0224)** | **0.089 (0.00109; 0.00163)** |
| ADHD | 0.081 | 0.079 | -0.007 (NS) | **0.091 (0.00146; 0.00293)** | **0.085 (0.00182; 0.00242)** |
| Schizophrenia | 0.273 | 0.352 | **0.103 (0.0198; 0.0396)** | **0.260 (4.46E-20; 5.35E-19)** | **0.342 (1.31E-36; 1.57E-35)** |
| Alcohol (drinks/wk) | 0.059 | 0.075 | 0.018 (NS) | **0.073 (0.0104; 0.0156)** | **0.084 (0.00207; 0.00248)** |
| Smoking (ever smoked) | 0.116 | 0.071 | -0.059 (NS) | **0.121 (2.49E-05; 7.48E-05)** | **0.076 (0.00602; 0.00657)** |
| Alzheimer's disease | -0.028 | 0.061 | **0.108 (0.0109; 0.0326)** | -0.026 (NS) | **0.064 (0.0193; 0.0193)** |

Notes. All tests were adjusted for sex, age, and 20 genetic ancestry principal components.
Results in bold type are significant.
All PGS were normalized to the UK controls, giving the UK controls mean PGS of zero.

### Sex-specific tests

Comparisons of SD-N vs. SD-S and to UK controls were done within sex, as shown in Table 3 and Figure 2a and 2b. Again, results were normalized to controls PRS. Results showed significantly lower PGS in the male SD-N vs. SD-S subtype comparisons for MDD and anxiety (FDR<0.05), and also for PTSD, neuroticism, and schizophrenia (FDR<0.10). In the male SD-N subtype, PGS for depressed affect, neuroticism, autism, and AD were not significantly different from controls. The PGS for smoking was elevated over controls only in SD-N, and the PGS for autism was only elevated over controls only in SD-S.

**Figure 2.**
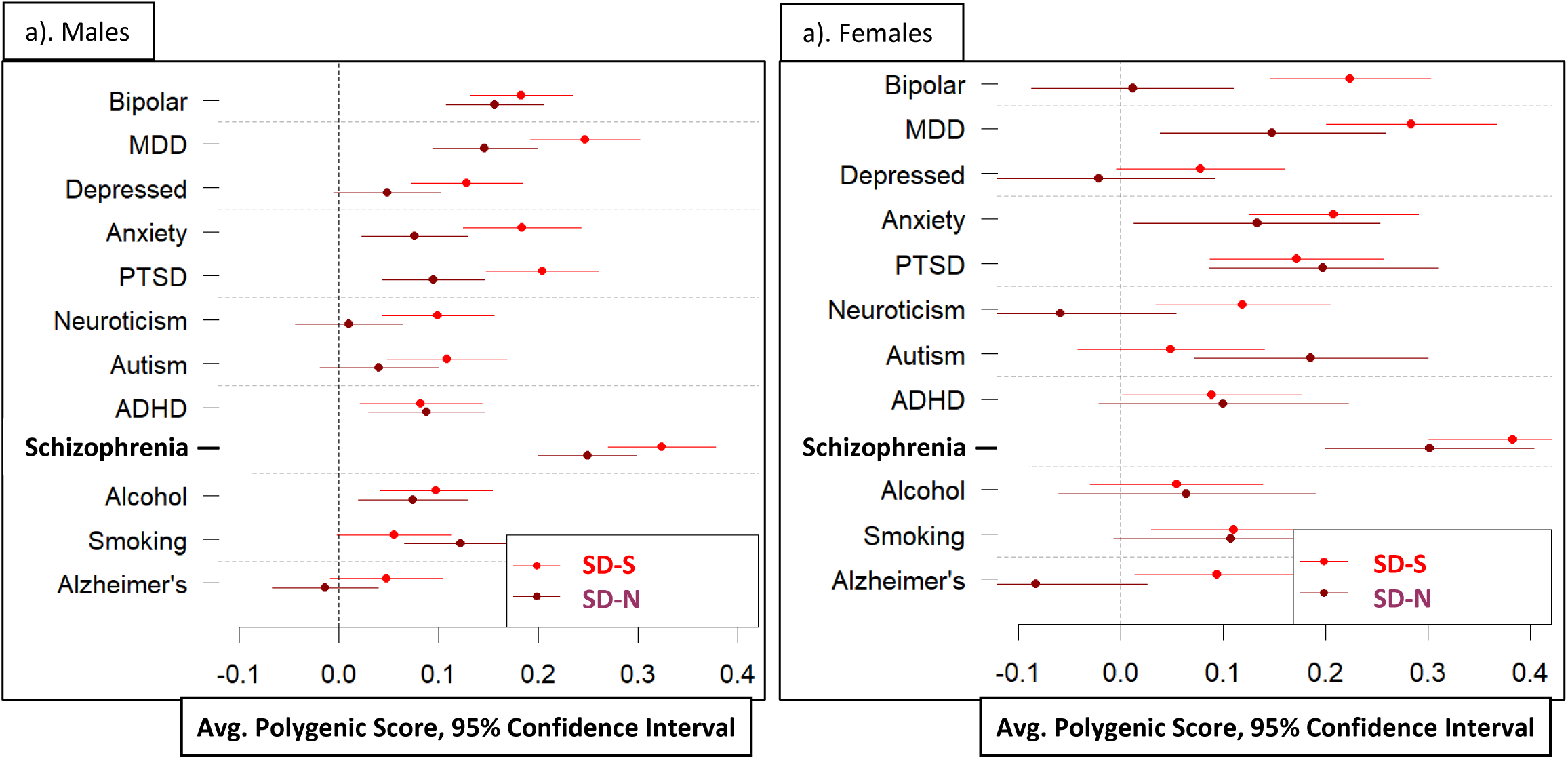
Polygenic score (PGS) analysis of covariance results for suicide mortality subtypes in males only (a) and females only (b). Results for suicide subtypes are normalized to the UK controls, which are represented by the zero lines in the graphs. All tests were adjusted for sex, age, and 20 genetic ancestry principal components. SD-N = suicide deaths with no evidence of prior nonfatal suicidality. SD-S = suicide deaths with evidence of prior nonfatal suicidality. MDD = major depressive disorder. PTSD = post-traumatic stress disorder. ADHD = attention deficit hyperactivity disorder.

**Table 3.** Sex-specific polygenic score (PGS) analysis of covariance results comparing suicide death subsets.

| Scored trait or diagnosis | SD-N mean PGS | SD-S mean PGS | SD-N vs. SD-S (p-value; FDR-adj p-value) | SD-N vs. UK Controls (p-value; FDR-adj p-value) | SD-S vs. UK Controls (p-value; FDR-adj p-value) |
| --- | --- | --- | --- | --- | --- |
| **MALES ONLY** |  |  |  |  |  |
| Bipolar disorder | 0.157 | 0.183 | 0.038 (NS) | **0.161 (5.32E-07; 3.20E-06)** | **0.187 (3.16E-08; 9.49E-08)** |
| MDD | 0.147 | 0.247 | **0.126 (0.0101; 0.0404)** | **0.150 (2.93E-06; 1.18E-05)** | **0.253 (9.34E-14; 5.61E-13)** |
| Depressed Affect | 0.048 | 0.128 | 0.097 (0.0433; 0.104) | 0.049 (NS) | **0.131 (1.09E-04; 1.87E-04)** |
| Anxiety | 0.076 | 0.183 | **0.126 (0.00781; 0.0469)** | **0.078 (0.0148; 0.0254)** | **0.186 (3.50E-08; 8.40E-08)** |
| PTSD | 0.095 | 0.204 | 0.136 (0.00514; 0.0617) | **0.097 (0.00240; 0.00576)** | **0.208 (7.683E-10; 3.07E-09)** |
| Neuroticism | 0.010 | 0.100 | 0.108 (0.0241; 0.0723) | 0.011 (NS) | **0.101 (0.00264; 0.00352)** |
| Autism | 0.041 | 0.109 | 0.070 (NS) | 0.041 (NS) | **0.109 (0.00113; 0.00200)** |
| ADHD | 0.050 | 0.164 | -0.006 (NS) | **0.088 (0.00533; 0.0107)** | **0.083 (0.0135; 0.0138)** |
| Schizophrenia | 0.249 | 0.324 | 0.103 (0.0441; 0.0883) | **0.255 (1.70E-15; 2.04E-14)** | **0.347 (2.11E-22; 2.53E-21)** |
| Alcohol (drinks/ week) | 0.074 | 0.098 | 0.028 (NS) | **0.075 (0.0180; 0.0270)** | **0.100 (0.00312; 0.00375)** |
| Smoking (ever smoked) | 0.122 | 0.056 | -0.075 (NS) | **0.124 (1.04E-04; 3.13E-04)** | 0.057 (NS) |
| Alzheimer's disease | -0.013 | 0.048 | 0.074 (NS) | 0.049 (NS) | -0.025 (NS) |
| **FEMALES ONLY** |  |  |  |  |  |
| Bipolar disorder | 0.012 | 0.224 | **0.300 (0.00151; 0.0181)** | 0.012 (NS) | **0.222 (1.85E-06; 7.38E-06)** |
| MDD | 0.148 | 0.283 | 0.166 (NS) | 0.149 (0.0225; 0.0654) | **0.285 (1.42E-09; 8.52E-09)** |
| Depressed Affect | -0.021 | 0.078 | 0.122 (NS) | -0.021 (NS) | 0.079 (NS) |
| Anxiety | 0.133 | 0.208 | 0.087 (NS) | 0.133 (0.0407; 0.0978) | **0.209 (8.91E-06; 2.67E-05)** |
| PTSD | 0.198 | 0.172 | -0.030 (NS) | **0.198 (0.00235; 0.0141)** | **0.172 (2.43E-04; 3.84E-04)** |
| Neuroticism | -0.059 | 0.119 | 0.207 (0.0156; 0.0624) | -0.059 (NS) | **0.119 (0.0111; 0.00184)** |
| Autism | 0.186 | 0.049 | -0.144 (NS) | **0.187 (0.00423; 0.0173)** | 0.049 (NS) |
| ADHD | 0.100 | 0.089 | -0.012 (NS) | 0.101 (NS) | 0.090 (NS) |
| Schizophrenia | 0.273 | 0.352 | 0.106 (NS) | **0.301 (3.66E-06; 4.33E-05)** | **0.381 (3.91E-16; 4.69E-15)** |
| Alcohol (drinks/ week) | 0.065 | 0.054 | -0.011 (NS) | 0.065 (NS) | 0.055 (NS) |
| Smoking (ever smoked) | 0.108 | 0.110 | 0.003 (NS) | 0.108 (NS) | **0.111 (0.0184; 0.00263)** |
| Alzheimer's disease | -0.082 | 0.094 | 0.229 (0.0114; 0.0686) | -0.083 (NS) | **0.094 (0.0444; 0.00555)** |

Note. All tests were adjusted for sex, age, and 20 genetic ancestry principal components.
Results in bold type are significant.
All PGS were normalized to the UK controls, giving the UK controls mean PGS of zero.
SD-N = suicide deaths with no evidence of prior nonfatal suicidality. SD-S = suicide deaths with evidence of prior nonfatal suicidality. MDD = major depressive disorder. PTSD = post-traumatic stress disorder. ADHD = attention deficit hyperactivity disorder. FDR-adj = adjusted for False Discovery Rate.

For females, the PGS was significantly lower in the SD-N vs. SD-S subtype test for bipolar disorder (FDR<0.05), and for neuroticism and AD (FDR<0.1). Compared to controls, the female SD-N subtype was significantly elevated for PTSD, schizophrenia, and autism (FDR<0.05), and for MDD and anxiety (FDR<0.1). The female SD-S subtype showed elevated PGS for all diagnoses except autism, ADHD, and depressed affect. Interestingly, the PGS for autism in females was elevated over controls for SD-N but not SD-S, opposite of the result for autism in males. For females, the smoking PGS was elevated over controls only in SD-S, again opposite the smoking result in males.

## DISCUSSION

Prior nonfatal suicidality is the most robust predictor of suicide mortality,^5–7^ but the association with this predictor is far from perfect, and a substantial proportion of suicide deaths occur in its absence.^8–10^ Previous work in the Utah Suicide Mortality Risk Study (USMRS)^15^ showed that suicides without prior nonfatal suicidality (SD-N) have significantly fewer diagnoses associated with neuropsychiatric conditions than those with prior nonfatal suicidality (SD-S). Prior thinking in suicidality research would place high likelihood that these differences would be due to less care-seeking or care access in the SD-N subtype, but assume that underlying liability to psychopathology would be a ubiquitous underlying factor in risk of suicide death.^29^ However, it is possible that suicide deaths without prior suicidality may have differences in underlying genetic liabilities to mental health. This study tests this hypothesis with polygenic scores associated with liabilities to neuropsychiatric conditions for 1,364 population-ascertained suicide deaths in the SD-N group. These suicide deaths were compared to 1,467 suicides with prior suicidality (SD-S). Evidence of suicidality included not only diagnostic codes, but also any mention of suicidal ideation or behaviors in clinical notes to ensure that the SD-N subtype was as accurate as possible. This study also included genetic data from 20,368 controls matched to the Utah suicides for genetic ancestry.

Demographic comparisons showed that the SD-N subtype was significantly younger and more male than the SD-S subtype; all analyses adjusted for these significant covariates. Polygenic scores (PGS) were tested from 12 common neuropsychiatric conditions with available summary statistics from recent published studies selected for size and similarity of genetic ancestry and ascertainment to the USMRS; this study design was preregistered to minimize bias.^16^

Results revealed significantly lower PGS (FDR<0.05) in SD-N vs. SD-S across the spectrum of neuropsychiatric conditions, including bipolar disorder, MDD, depressed affect, anxiety, PTSD, neuroticism, schizophrenia, and AD. In addition, for depressed affect, neuroticism, and AD, the SD-N subtype was not significantly different from the controls. These results indicate a substantially lower degree of underlying genetic liability to many aspects of mental health in the SD-N subtype.

Results have implications for future research and also clinical interventions; namely, all suicides do not have similar elevation of underlying liability to neuropsychiatric conditions. For research, combining suicide deaths with and without evidence of prior nonfatal suicidality will mix two substantially different genetic subtypes, therefore weakening results specific to suicide mortality. Regarding clinical interventions, results suggest a focus solely on mental health risks when designing screening and interventions may be less effective for those for whom this underlying liability may be substantially attenuated. Further understanding of both clinical characteristics and underlying genetic liabilities in SD-N will likely be required to direct more targeted, effective screening and interventions.

Our analyses also indicated that while many conditions showed attenuation of PGS in SD-N, PGS for ADHD and alcohol were instead equally elevated over controls in both suicide subtypes. These results suggest these underlying genetic liabilities, and behaviors associated with them such as poor impulse regulation,^30,31^ may be common to suicide mortality risk regardless of the presence of prior suicidality. PGS for autism and smoking were also not attenuated in SD-N vs. SD-S in the combined analysis. However, sex-specific results suggested a potentially more complex picture for these traits. For smoking, PGS was elevated only in SD-S vs. controls in females, suggesting aspects of smoking PGS risk in female suicide may be more closely associated with genetic risks of psychopathology. In men, the PGS for smoking was elevated over controls only in SD-N, suggesting non-psychiatric aspects of the smoking PGS may be more tied to risk of suicide mortality in men. For autism, PGS was elevated in males only for SD-S, suggesting that for males with neurodevelopmental genetic risk, co-occurring psychopathology may be important for risk of suicide death. Conversely, for females, autism PGS was elevated only in the SD-N subset, suggesting studies in females with neurodevelopmental risks should perhaps address a broader range of characteristics beyond psychopathology (e.g., physical health liabilities, or basic behavioral traits associated with autism, such as cognitive rigidity or social and communication deficits).^32^ Other sex differences suggested that males may drive the overall attenuation of PGS in SD-N for MDD and anxiety, and also to a lesser degree, PTSD and schizophrenia. In contrast, lower PGS in SD-N for bipolar disorder, AD, and, to a lesser extent, neuroticism may be more driven by females.

Finally, the PGS showing the most significant elevation over controls was schizophrenia, and while there was substantial attenuation in SD-N vs. SD-S, PGS for schizophrenia in SD-N still showed the largest elevation over controls of any of the tested traits. Interestingly, in previously studied clinical data in these subsets in our data resource, rates of overt schizophrenia diagnoses in SD-S were high (12.8%), but for SD-N rates of clinically diagnosed schizophrenia were only modestly elevated over controls who had no evidence of lifetime suicidality (1.5%).^15^ The clinical data therefore did not indicate a high rate of overt schizophrenia in the SD-N subtype. While it is possible that the SD-N group may have some individuals with undiagnosed schizophrenia, it is also possible this PGS elevation in SD-N represents additional sensitivity of the schizophrenia PGS to detect significant subclinical liability, or that the elevation represents other aspects of physical or behavioral traits included in the score reflecting genetic liability to schizophrenia^33,34^ that are associated with suicide mortality.

Overall, the results suggest that pooling PGS across sex may dampen statistical signals of risk, especially when associations show effects in opposite directions by sex (e.g.,, smoking and autism). Sex-specific findings suggest that there may be underlying genetic liabilities leading to suicide mortality that differ substantially by sex for certain clinical mental health outcomes. Future genetic liability risk discovery efforts investigating both psychiatric and non-psychiatric outcomes leading to suicide mortality should stratify both by presence of prior nonfatal suicidality and also by sex.

### Limitations

While the NLP of all clinical notes to identify suicidality greatly improved the capture of individuals in the SD-S group, it remains possible that some in the SD-N group had prior suicidality not represented either in diagnostic codes or the NLP. However, such individuals would more closely resemble the SD-S group, resulting in more conservative SD-N vs. SD-S outcomes. In addition, controls are unscreened for suicidality, again resulting in potential lessening of differences between controls and suicide subsets. The USMRS study, while population-ascertained, has individuals of predominantly European ancestry, limiting generalizability. Sample sizes of each suicide death subset are relatively small, requiring replication and extension with future larger data resources. Small samples were particularly apparent for sex-specific analyses, where results should be interpreted with caution. Our study also included only pre-registered PGS associated with psychopathology to control for bias. A more general screen of PGS across additional psychiatric and medical conditions in these interesting suicide death subsets is warranted. In addition, while our results stratified by sex, it is likely that other demographic stratifications may also reveal important results. Finally, our study did not address mediating or moderating effects of social and environmental exposures, which will be critical to achieve a fuller understanding of mortality risk.

### Conclusions

While these limitations suggest our knowledge of risks leading to suicide mortality is far from complete, this study provides important first steps in determining underlying risks in suicide deaths with no evidence of prior nonfatal suicidality, a large subtype previously unavailable for study. Results of this work show that previously demonstrated dramatically reduced evidence of neuropsychiatric diagnoses in clinical data in this suicide subset is due, at least in part, to attenuation of underlying polygenic liabilities associated with neuropsychiatric conditions rather than simply a lack of access to care. Results also justify additional studies of non-psychiatric characteristics, and indicate that future studies within sex will be essential. In summary, suicide deaths without evidence of prior nonfatal suicidality, who represent roughly half of those who die, appear to have significantly different genetic etiology from suicide deaths who do have the robust risk predictor of prior suicidality. These findings have far-reaching implications both for future research and for identification and intervention for those at highest risk of mortality.

## Data Availability

All data produced in the present study are available upon request to the authors with the additional required approvals for disclosure of the data by the Utah Department of Health and Human Services and the University of Utah.

## ACKNOWLEDGMENTS

This work was supported by the National Institute of Mental Health (HC, grant number R01MH122412, R01MH123489; AVB, grant number R01ES032028; AD, grant number R01MH123619), a research contract from Janssen Research & Development, LLC (HC, QL); the American Foundation for Suicide Prevention (ED, grant number BSG-1-005-18), the Brain & Behavior Research Foundation--NARSAD (ED, grant number 28132; AS grant number 28686; EM grant number 31248); and the Clark Tanner Foundation (HC, AS, EM, AVB). We thank the Utah Population Database (UPDB) staff of the Huntsman Cancer Institute, University of Utah (funded in part by the Huntsman Cancer Foundation) for their role in the ongoing collection, maintenance, and support of the UPDB, and we acknowledge partial support of the UPDB through grant P30 CA2014 from the National Cancer Institute and through the Huntsman Cancer Institute. We thank University of Utah Health Data Science Services for data and analytics support, and the University of Utah Pedigree and Population Resource and the University of Utah Health Enterprise Data Warehouse for establishing the Master Subject Index between the Utah Population Database and the University of Utah Health Sciences Center (NCRR, R01RR021746) with additional support from the Utah Department of Health and Human Services. We also thank data experts at the Intermountain Data Warehouse particularly for their assistance with data linking and with the natural language processing pipeline used for this study. DNA extraction from Utah suicide deaths was performed by the University of Utah Center for Clinical and Translational Science supported by the National Center for Advancing Translational Sciences of the NIH (grant number UL1TR002538). Genotyping of Utah suicide deaths was performed by the University of Utah Genomic Core (UL1TR002538) and by Illumina, Inc. with support from Janssen Research & Development, LLC. The support and resources from the Center for High Performance Computing at the University of Utah are also gratefully acknowledged. Generation Scotland received core support from the Chief Scientist Office of the Scottish Government Health Directorates [CZD/16/6] and the Scottish Funding Council [HR03006] and is currently supported by the Wellcome Trust [216767/Z/19/Z]. Genotyping of the GS:SFHS samples was carried out by the Genetics Core Laboratory at the Edinburgh Clinical Research Facility, University of Edinburgh, Scotland and was funded by the Medical Research Council UK and the Wellcome Trust (Wellcome Trust Strategic Award “STratifying Resilience and Depression Longitudinally” (STRADL) Reference 104036/Z/14/Z).

## Conflicts

Dr. Qingqin Li previously was employed in the Neuroscience Therapeutic Area, Janssen Research and Development, Titusville, NJ. She is now Scientific Director of Human Genetics / Genomics at CHDI, Cure Huntington’s Disease Initiative.

## Notes

### Author Declarations

This study was approved by Institutional Review Boards from the University of Utah, Intermountain Health, and the Utah Department of Health and Human Services.

## REFERENCES

1. Curtin MA, Garnett MF, Ahmad FB. Provisional estimates of suicide by demographic characteristics: United States, 2022. NVSS Vital Statistics Rapid Release, Report No. 34, Nov 2023. Centers for Disease Control and Prevention, National Center for Health Statistics.

2. World Health Organization. Suicide fact sheet. 2024 29 Aug, https://www.who.int/news-room/fact-sheets/detail/suicide.

3. Belsher BE, Smolenski DJ, Pruitt LD, Bush NE, Beech EH, Workman DE, Morgan RL, Evatt DP, Tucker J, Skopp NA. Prediction Models for Suicide Attempts and Deaths: A Systematic Review and Simulation. JAMA Psychiatry. 2019 Jun 1;76(6):642–651.

4. Franklin JC, Ribeiro JD, Fox KR, Bentley KH, Kleiman EM, Huang X, Musacchio KM, Jaroszewski AC, Chang BP, Nock MK. Risk factors for suicidal thoughts and behaviors: A meta-analysis of 50 years of research. Psychol Bull. 2017 143;187–232.

5. Liu BP, Lunde KB, Jia CX, Qin P. The short-term rate of non-fatal and fatal repetition of deliberate self-harm: A systematic review and meta-analysis of longitudinal studies. J Affect Disord. 2020 Aug 1;273:597–603.

6. Carroll R, Metcalfe C, Gunnell D. Hospital presenting self-harm and risk of fatal and non-fatal repetition: systematic review and meta-analysis. PLoS One. 2014 Feb 28;9(2):e89944.

7. Owens D, Horrocks J, House A. Fatal and non-fatal repetition of self-harm: systematic review. Br J Psychiatry. 2002;181:193–199.

8. Jordan JT, McNiel DE. Characteristics of persons who die on their first suicide attempt: results from the National Violent Death Reporting System. Psychol Med. 2020 Jun;50(8):1390–1397.

9. Botswick JM, Pabbati CJ, Geske JR, McKean AJ. Suicide Attempt as a Risk Factor for Completed Suicide: Even More Lethal Than We Knew. Am J Psychiatry 2016, 173(11), 1094–1100.

10. Isometsä ET, Lönnqvist JK. Suicide attempts preceding completed suicide. Br J Psychiatry. 1998 Dec;173:531–535.

11. Xiao Y, Bi K, Yip PS, Cerel J, Brown TT, Peng Y, Pathak J, Mann JJ. Decoding suicide decedent profiles and signs of suicidal intent using latent class analysis. JAMA Psychiatry. 2024 Mar 20.

12. Stone DM, Simon TR, Fowler KA, et al. Vital signs: Trends in state suicide rates—United States, 1999-2016 and circumstances contributing to suicide—27 states, 2015. MMWR Morb Mortal Wkly Rep. 2018 Jun 8;67:617–624.

13. Stene-Larsen K, Reneflot A. Contact with primary and mental health care prior to suicide: a systematic review of the literature from 2000 to 2017. Scand J Pub Health 2019 47:9–17.

14. Benjamini Y, Hochberg Y. Controlling the false discovery rate: A practical and powerful approach to multiple testing. J Royal Statistical Soc B (Methodological). 1995, 57(1):289–300.

15. Coon H, Shabalin A, DiBlasi E, Monson ET, Han S, Kaufman EA, Chen D, Kious B, Molina N, Yu Z, Staley M, Crockett DK, Colbert SM, Mullins N, Bakian AV, Docherty AR, Keeshin B. Absence of nonfatal suicidal behavior preceding suicide death reveals differences in clinical risks. Psychiatry Res. 2025 Feb 24;347:116391. Epub ahead of print.

16. Open Science Framework Registry public study design registration created 2024, 5 Aug, 10.17605/OSF.IO/KCYWG

17. Coon H, Shabalin A, Bakian AV, DiBlasi E, Monson ET, Kirby A, Chen D, Fraser A, Yu Z, Staley M, Callor WB, Christensen ED, Crowell SE, Gray D, Crockett DK, Li QS, Keeshin B, Docherty AR. Extended familial risk of suicide death is associated with younger age at death and elevated polygenic risk of suicide. Am J Med Genet B Neuropsychiatr Genet. 2022 Apr;189(3-4):60–73.

18. Docherty AR, Shabalin AA, DiBlasi E, Monson E, Mullins N, Adkins DE, Bacanu SA, Bakian AV, Crowell S, Chen D, Darlington TM, Callor WB, Christensen ED, Gray D, Keeshin B, Klein M, Anderson JS, Jerominski L, Hayward C, Porteous DJ, McIntosh A, Li Q, Coon H. Genome-wide association study of suicide death and polygenic prediction of clinical antecedents. Am J Psychiatry. 2020 Oct 1;177(10):917–927.

19. Utah Population Database. Database overview. 2024 Aug 15. https://uofuhealth.utah.edu/huntsman/utah-population-database/data

20. UK10K Consortium; Walter K, Min JL, Huang J, Crooks L, Memari Y, McCarthy S, Perry JR, Xu C, Futema M, Lawson D, Iotchkova V, Schiffels S, Hendricks AE, Danecek P, Li R, Floyd J, Wain LV, Barroso I, Humphries SE, Hurles ME, Zeggini E, Barrett JC, Plagnol V, Richards JB, Greenwood CM, Timpson NJ, Durbin R, Soranzo N. The UK10K project identifies rare variants in health and disease. Nature. 2015 Oct 1;526(7571):82–90.

21. Smith BH, Campbell H, Blackwood D, Connell J, Connor M, Deary IJ, Dominiczak AF, Fitzpatrick B, Ford I, Jackson C, Haddow G, Kerr S, Lindsay R, McGilchrist M, Morton R, Murray G, Palmer CN, Pell JP, Ralston SH, St Clair D, Sullivan F, Watt G, Wolf R, Wright A, Porteous D, Morris AD. Generation Scotland: the Scottish Family Health Study; a new resource for researching genes and heritability. BMC Med Genet. 2006 Oct 2;7:74.

22. Hedegaard H, Schoenbaum M, Claassen C, Crosby A, Holland K, Proescholdbell S. Issues in Developing a Surveillance Case Definition for Nonfatal Suicide Attempt and Intentional Self-harm Using International Classification of Diseases, Tenth Revision, Clinical Modification (ICD-10-CM) Coded Data. Natl Health Stat Report. 2018 Feb;(108):1–19. PMID: 29616901.

23. Illumina Research Group. Infinium PsychArray-24 (24-sample). Illumina.com, 2025. https://emea.illumina.com/library-prep-array-kit-selector/kits-and-arrays/psycharray_24-sample.html#:~:text=The%20Infinium%20PsychArray%2D24%20BeadChip,on%20psychiatric%20predisposition%20and%20risk.

24. Illumina Research Group. GenomeStudio Software. Illumina.com, 2025. https://www.illumina.com/techniques/microarrays/array-data-analysis-experimentaldesign/genomestudio.html

25. Genovese G. kgp2anc: A protocol to estimate ancestry from the main ethnicities (European, African, East Asian, Native American, South Asian) using principal components analysis together with individuals from the 1000 Genomes project phase 3 starting from raw Illumina genotype array data. Github 2025, code and documentation at https://github.com/freeseek/kgp2anc.

26. The 1000 Genomes Project Consortium. A global reference for human genetic variation. Nature 526, 68–74 (2015).

27. Choi SW, O’Reilly PF. PRSice-2: Polygenic Risk Score software for biobank-scale data. Gigascience. 2019 Jul 1;8(7):giz082.

28. Fritsche LG, Gruber SB, Wu Z, Schmidt EM, Zawistowski M, Moser SE, Blanc VM, Brummett CM, Kheterpal S, Abecasis GR, Mukherjee B. Association of Polygenic Risk Scores for Multiple Cancers in a Phenome-wide Study: Results from The Michigan Genomics Initiative. Am J Hum Genet. 2018 Jun 7;102(6):1048–1061.

29. Chesney E, Goodwin GM, Fazel S. Risks of all-cause and suicide mortality in mental disorders: a meta-review. World Psychiatry. 2014;13(2):153–60.

30. Koirala S, Grimsrud G, Mooney MA, Larsen B, Feczko E, Elison JT, Nelson SM, Nigg JT, Tervo-Clemmens B, Fair DA. Neurobiology of attention-deficit hyperactivity disorder: historical challenges and emerging frontiers. Nat Rev Neurosci. 2024 Dec;25(12):759–775.

31. Lomas C. Neurobiology, psychotherapeutic interventions, and emerging therapies in addiction: a systematic review. J Addict Dis. 2024 Dec 17:1–19.

32. Happé F, Ronald A, Plomin R. Time to give up on a single explanation for autism. Nat Neurosci. 2006 Oct;9(10):1218–20.

33. Szoke A, Pignon B, Godin O, Ferchiou A, Tamouza R, Leboyer M, Schürhoff F. Multimorbidity and the Etiology of Schizophrenia. Curr Psychiatry Rep. 2024 May;26(5):253–263.

34. Romero C, de Leeuw C, Schipper M, Maciel BAPC, van den Heuvel MP, Brouwer RM, Smit AB, Koopmans F, Posthuma D, van der Sluis S. Immune-developmental processes contribute to schizophrenia risk: insights from a genetic overlap study with height. Biol Psychiatry. 2025 Mar 10:S0006-3223(25)01018–2.

35. Mullins N, Forstner AJ, O’Connell KS, Coombes B, Coleman JRI, Qiao Z, Als TD, et al. Genome-wide association study of more than 40,000 bipolar disorder cases provides new insights into the underlying biology. Nat Genet. 2021 Jun;53(6):817–829.

36. Als TD, Kurki MI, Grove J, Voloudakis G, Therrien K, Tasanko E, Nielsen TT, et al. Depression pathophysiology, risk prediction of recurrence and comorbid psychiatric disorders using genome-wide analyses. Nat Med. 2023 Jul;29(7):1832–1844.

37. Nagel M, Jansen PR, Stringer S, Watanabe K, de Leeuw CA, Bryois J, Savage JE, et al. Meta-analysis of genome-wide association studies for neuroticism in 449,484 individuals identifies novel genetic loci and pathways. Nat Genet. 2018 Jul;50(7):920–927.

38. Meier SM, Trontti K, Purves KL, Als TD, Grove J, Laine M, Pedersen MG, et al. Genetic Variants Associated With Anxiety and Stress-Related Disorders: A Genome-Wide Association Study and Mouse-Model Study. JAMA Psychiatry. 2019 Sep 1;76(9):924–932.

39. Nievergelt CM, Maihofer AX, Klengel T, Atkinson EG, Chen CY, Choi KW, Coleman JRI, et al. International meta-analysis of PTSD genome-wide association studies identifies sex- and ancestry-specific genetic risk loci. Nat Commun. 2019 Oct 8;10(1):4558.

40. Trubetskoy V, Pardiñas AF, Qi T, Panagiotaropoulou G, Awasthi S, Bigdeli TB, Bryois J, …, Schizophrenia Working Group of the Psychiatric Genomics Consortium. Mapping genomic loci implicates genes and synaptic biology in schizophrenia. Nature. 2022 Apr;604(7906):502–508.

41. Grove J, Ripke S, Als TD, Mattheisen M, Walters RK, Won H, Pallesen J, et al. Identification of common genetic risk variants for autism spectrum disorder. Nat Genet. 2019 Mar;51(3):431–444.

42. Demontis D, Walters RK, Martin J, Mattheisen M, Als TD, Agerbo E, Baldursson G, et al. Discovery of the first genome-wide significant risk loci for attention deficit/hyperactivity disorder. Nat Genet. 2019 Jan;51(1):63–75.

43. Karlsson Linnér R, Biroli P, Kong E, Meddens SFW, Wedow R, Fontana MA, Lebreton M, et al. Genome-wide association analyses of risk tolerance and risky behaviors in over 1 million individuals identify hundreds of loci and shared genetic influences. Nat Genet. 2019 Feb;51(2):245–257.

44. Wightman DP, Jansen IE, Savage JE, Shadrin AA, Bahrami S, Holland D, Rongve A, et al. A genome-wide association study with 1,126,563 individuals identifies new risk loci for Alzheimer’s disease. Nat Genet. 2021 Sep;53(9):1276–1282.

