## Supplementary Table S1 for "Different genetic liabilities to neuropsychiatric conditions in suicides with no prior suicidality"

Supplementary Table S1. Source studies for polygenic score statistics used to test for underlying genetic vulnerabilities.

| **Phenotype (source study)** | **N source study participants** | **Notes** |
| --- | --- | --- |
| Bipolar Disorder^35^ | 41.917 BD; 371,549 controls | Lifetime BD (ICD-9/ICD-10 diagnoses; EUR) |
| Major Depressive Disorder^36^ | 30,618 MDD; 38,200 controls | Lifetime MDD (ICD-10 diagnoses; EUR) |
| Depressed Affect^37^ | 357,957 depression spectrum symptoms | UKBiobank, Genetics of Personality Consortium (questionnaire, EUR) |
| Neuroticism^37^ | 390,278 (quantitative neuroticism symptoms) | UKBiobank, Genetics of Personality Consortium (questionnaire, EUR) |
| Anxiety^38^ | 12,655 anxiety disorders; 19,225 controls | iPsych, Comprehensive anxiety diagnoses from ICD-10 (GAD, agoraphobia, panic, phobias, mixed anxiety disorder; EUR) |
| PTSD^39^ | 32,428 PTSD; 174,227 controls | EUR subset of meta-analysis of European research resources, non-military ascertainment (multiple diagnostic methods; EUR) |
| Schizophrenia^40^ | 67,390 SZ; 94,015 controls | SZ, Psychiatric Genetics Consortium and other worldwide data (multiple diagnostic methods; EUR) |
| Autism^41^ | 18,381 autism; 27,969 controls | Lifetime autism spectrum disorder (ICD-10; EUR) |
| ADHD^42^ | 19,099 ADHD; 34,194 controls | Lifetime ADHD (ICD-10 diagnoses; EUR) |
| Alcohol^43^ | 414,343 individuals (quantitative data) | Drinks per week (UKBiobank, questionnaire; EUR) |
| Smoking^43^ | 518,633 individuals | Lifetime ever smoked (UKBiobank, questionnaire; EUR) |
| Alzheimer’s Disease^44^ | 90,338 AD cases; 1,036,225 controls | AD diagnoses, UKBiobank and 13 other studies (multiple diagnostic methods; EUR) |

Notes. BD = Bipolar Disorder; MDD = Major Depressive Disorder; PTSD = Post-Traumatic Stress Disorder; ADHD = Attention Deficit Hyperactivity Disorder; GAD = Generalized Anxiety Disorder; AD = Alzheimer’s Disease. EUR = European genetic ancestry. ICD = International Classification of Diseases.
